# Association of *HLA-class II* alleles with risk of relapse in myeloperoxidase-antineutrophil cytoplasmic antibody positive vasculitis in the Japanese population

**DOI:** 10.1101/2022.12.28.22283983

**Authors:** Aya Kawasaki, Ken-ei Sada, Premita Ari Kusumawati, Fumio Hirano, Shigeto Kobayashi, Kenji Nagasaka, Takahiko Sugihara, Nobuyuki Ono, Takashi Fujimoto, Makio Kusaoi, Naoto Tamura, Yasuyoshi Kusanagi, Kenji Itoh, Takayuki Sumida, Kunihiro Yamagata, Hiroshi Hashimoto, Hirofumi Makino, Yoshihiro Arimura, Masayoshi Harigai, Naoyuki Tsuchiya

**Author notes:** **Corresponding authors:** Aya Kawasaki, Naoyuki Tsuchiya.

## Abstract

**Background:** Disease relapse remains a major problem in the management of anti-neutrophil cytoplasmic antibody (ANCA)-associated vasculitis (AAV). In European populations, *HLA-DPB1*04:01* is associated with both susceptibility and relapse risk in proteinase 3-ANCA positive AAV. In a Japanese population, we previously reported an association between *HLA-DRB1*09:01* and *DQB1*03:03* with susceptibility to, and *DRB1*13:02* with protection from, myeloperoxidase-ANCA positive AAV (MPO-AAV). Subsequently, the association of *DQA1*03:02*, which is in strong linkage disequilibrium with *DRB1*09:01* and *DQB1*03:03*, with MPO-AAV susceptibility was reported in a Chinese population. However, an association between these alleles and risk of relapse has not yet been reported. Here, we examined whether *HLA-class II* is associated with the risk of relapse in MPO-AAV.

**Methods:** First, the association of *HLA-DQA1*03:02* with susceptibility to MPO-AAV and microscopic polyangiitis (MPA) and its relationship with previously reported *DRB1*09:01* and *DQB1*03:03* were examined in 440 Japanese patients and 779 healthy controls. Next, the association with risk of relapse was analyzed in 199 MPO-ANCA positive, PR3-ANCA negative patients enrolled in previously reported cohort studies on remission induction therapy. Uncorrected P values (P_uncorr_) were corrected for multiple comparisons in each analysis using the false discovery rate method.

**Results:** The association of *DQA1*03:02* with susceptibility to MPO-AAV and MPA was confirmed in a Japanese population (MPO-AAV: P_uncorr_=5.8×10^−7^, odds ratio [OR] 1.74, 95% confidence interval [CI] 1.40–2.16, MPA: P_uncorr_=1.1×10^−5^, OR 1.71, 95%CI 1.34–2.17). *DQA1*03:02* was in strong linkage disequilibrium with *DRB1*09:01* and *DQB1*03:03*, and the causal allele could not be determined using conditional logistic regression analysis. Relapse-free survival was shorter with nominal significance in carriers of *DRB1*09:01* (P_uncorr_=0.049, Q=0.42, hazard ratio [HR]:1.87), *DQA1*03:02* (P_uncorr_=0.020, Q=0.22, HR:2.11) and *DQB1*03:03* (P_uncorr_=0.043, Q=0.48, HR:1.91) than in non-carriers in the log-rank test. Conversely, serine carriers at position 13 of HLA-DRβ1 (HLA-DRβ1_13S), including *DRB1*13:02* carriers, showed longer relapse-free survival with nominal significance (P_uncorr_=0.010, Q=0.42, HR:0.31). By combining *DQA1*03:02* and HLA-DRβ1_13S, a significant difference was detected between groups with the highest and lowest risk for relapse (P_uncorr_=0.0055, Q=0.033, HR:4.02).

**Conclusion:** *HLA-class II* is associated not only with susceptibility to MPO-AAV but also with risk of relapse in the Japanese population.

## Introduction

Anti-neutrophil cytoplasmic antibody (ANCA)-associated vasculitis (AAV) is a group of necrotizing small vessel vasculitides characterized by ANCA production, mainly against proteinase 3 (PR3) or myeloperoxidase (MPO). AAV is classified as microscopic polyangiitis (MPA), granulomatosis with polyangiitis (GPA), or eosinophilic granulomatosis with polyangiitis (EGPA) according to the European Medicines Agency (EMA) algorithm (1). In AAV, epidemiological differences between European and Asian populations are well known. In European populations, GPA and PR3-ANCA positive AAV (PR3-AAV) are predominant, whereas in the Japanese population, MPA and MPO-ANCA-positive AAV (MPO-AAV) account for the majority of AAV cases (2). Such ethnic differences in epidemiology imply that the genetic background of AAV may play a role in its development.

Although patients with AAV achieve remission with immunosuppressive therapy (3), a substantial proportion experience relapse. In European populations, relapse occurs in approximately half of the patients with GPA within 5 years after achieving complete remission (4). Among AAV patients, the risk of relapse has been shown to be higher in those with GPA and PR3-AAV compared with MPA and MPO-AAV (5). In Japanese observational studies conducted to identify risk factors for AAV relapse, the dosage and tapering speed of prednisolone were associated with relapse (6, 7).

With respect to susceptibility to MPA and GPA, three genome-wide association studies (GWAS) in European populations have been reported thus far (8-10). In GPA and PR3-AAV, the most striking association was identified in the *HLA-DP* region, which is consistent with a previously reported association of GPA with *HLA-DPB1*04:01* in a German population (11). Additionally, *PRTN3* and *SERPINA1* genes, encoding PR3 and α1-antitrypsin, respectively, were identified as susceptibility genes (8,10). With respect to MPA and MPO-AAV, the *HLA-DQ* region has been associated with susceptibility in the GWAS (8,10). In agreement with this, the *HLA-DRB1*09:01*-*DQB1*03:03* haplotype was found to be associated with susceptibility to MPA and MPO-AAV in the Japanese population (12-15). In addition, the *DQA1*03:02-DQB1*03:03* haplotype was recently reported to be associated with MPO-AAV in a Chinese population (16). The *HLA-DRB1*09:01*-*DQA1*03:02*-*DQB1*03:03* haplotype is common in genera East Asian populations, but rare in European populations. Additionally, *DRB1*13:02* has been found to be associated with protection from MPA and MPO-AAV in the Japanese population (14).

Although several genes are associated with AAV susceptibility, those associated with relapse have not been well-characterized. In European populations, *HLA-DPB1*04:01*, the susceptibility allele for GPA, has been shown to be associated with a higher risk of AAV (17) and PR3-AAV relapse (18). However, similar studies have not been conducted in East Asian populations, in which MPA and MPO-AAV account for the majority of AAV cases.

In this study, we focused on MPO-AAV in the Japanese population, and examined whether *HLA-DRB1, DQA1, DQB1* and *DPB1* alleles are also associated with relapse in MPO-AAV, based on clinical data from Japanese nationwide cohort studies on remission induction therapy in AAV. Data were obtained from “Remission Induction Therapy in Japanese Patients with ANCA-associated Vasculitides” (RemIT-JAV), registered with the University Hospital Medical Information Network Clinical Trials Registry (UMIN000001648) (19) and “Remission Induction Therapy in Japanese Patients with ANCA-associated Vasculitides and Rapidly Progressive Glomerulonephritis” (RemIT-JAV-RPGN) (UMIN000005136) (20) carried out under the initiatives of Japan Research Committee of the Ministry of Health, Labour, and Welfare for Intractable Vasculitis (JPVAS) and Research Committee of Intractable Renal Disease of the Ministry of Health, Labour, and Welfare of Japan.

## Materials and Methods

### Patients and controls

We first examined whether *HLA-DQA1*03:02*, which has recently been reported in a Chinese population (16), is also associated with susceptibility to MPO-AAV and MPA in the Japanese population, and investigated its relationship with *DRB1*09:01* and *DQB1*03:03* (12-15), in 440 patients with AAV (male: 168, female: 272) and 779 controls (male: 306, female: 473) The breakdown is shown in Table 1. Among these subjects, 362 with MPO-AAV, 273 with MPA patients and 514 controls were included in *HLA-DRB1* and *DPB1* analyses in our previous study (14).

**Table 1.** Characteristics of the patients and healthy controls examined for the association of *HLA-DRB1*09:01* and *DQA1*03:02* with susceptibility to MPO-AAV and MPA.

|  | MPO-AAV/MPA | Healthy controls |
| --- | --- | --- |
| n | 440 | 779 |
| Sex (Male : Female) | 168 : 272 | 306 : 473 |
| EMA classification |  |  |
| MPA | 305 | NA |
| GPA | 64 | NA |
| EGPA | 36 | NA |
| Unclassifiable | 35 | NA |
| ANCA specificity |  |  |
| MPO-ANCA | 428 | NA |
| PR3-ANCA | 18 | NA |

n, number; SD, standard deviation; MPO-AAV, MPO-ANCA positive ANCA-associated vasculitis; MPA, microscopic polyangiitis; GPA, granulomatosis with polyangiitis; EGPA, eosinophilic granulomatosis with polyangiitis; NA, not applicable. Note that MPO-ANCA negative GPA, EGPA and unclassifiable AAV are not included in this study. Detailed information is available in Supplementary Data 1.

Genomic DNA samples were obtained from institutes participating in Japan Research Committee of the Ministry of Health, Labour, and Welfare for Intractable Vasculitis (JPVAS) and the Research Committee of Progressive Renal Disease, both organized by the Ministry of Health, Labour, and Welfare of Japan, as well as from research groups organized by Tokyo Medical and Dental University and University of Tsukuba. A total of 264 control samples were obtained from the Health Science Research Resources Bank (Osaka, Japan).

Among patients with MPO-AAV, 88 enrolled in RemIT-JAV (19) and 176 enrolled in RemIT-JAV-RPGN (20) were analyzed for relapse-free survival. The characteristics of these cohorts have been previously described (19,20). Briefly, the enrollment period was from April 2009 to December 2010 (RemIT-JAV) and from April 2011 to March 2014 (RemIT-JAV-RPGN). The enrollment criteria were as follows: 1) receiving a diagnosis of AAV by site investigators, 2) fulfilling the criteria for primary systemic vasculitis as proposed by the EMA algorithm, and 3) starting immunosuppressive treatment based on the discretion of the site investigators. Among these patients, 214 were MPO-ANCA positive and PR3-ANCA negative (thereafter, “MPO-ANCA single-positive”). To examine the rate of relapse after remission, 199 patients with MPO-ANCA single-positive AAV who achieved remission during the observation period were analyzed in this study. The observation period was 730 d after treatment initiation. Of the 199 patients included in this study, 8 died during the observation period (3 after relapse, and 5 without relapse). The causes of death in patients who died without relapse were infection (n=3), heart failure (n=1) and malignancy (n=1). These five patients were censored on the date of death. Remission was defined based on the Birmingham vasculitis activity score 2003 (21) of zero on two consecutive occasions at least one month apart (22). Relapse was defined as the recurrence or new onset of clinical signs and symptoms attributable to active vasculitis (6, 23).

Detailed information on the subjects is provided in Supplementary Data 1 (https://doi.org/10.6084/m9.figshare.21876159.v3).

### Genotyping

In all patients and controls, *HLA-DRB1* alleles were determined at the four-digit level using a WAKFlow HLA typing kit (Wakunaga Pharmaceutical Co., Ltd., Osaka, Japan) based on polymerase chain reaction-sequence-specific oligonucleotide probes (PCR-SSOP). *HLA-DQA1, DQB1* and *DPB1* alleles were genotyped using this system for the199 patients with MPO-AAV examined for relapse-free survival,

In susceptibility analysis, rs11545686C was used as a proxy for *HLA-DQA1*03:02*. Genotyping of rs11545686 was conducted by Sanger sequencing using 3130xl Genetic Analyzer (Thermo Fisher Scientific, Waltham, MA, USA). For amplification of *HLA-DQA1* region surrounding rs11545686, forward (5’-TTTGGTTTGGGTGTCTTCAGATT-3’) and reverse (5’-AAAGTTGTTCAGGGAAATTTGAGAATG-3’) primers (Primer ID: Hs00412887_CE, Thermo Fisher Scientific) were used and cycle sequencing reaction was conducted using the forward primer and the BigDye Terminator v3.1 Cycle Sequencing Kit (Thermo Fisher Scientific). Representative Sanger sequencing chromatograms for each genotype are shown in Supplementary Figure S1 (https://doi.org/10.6084/m9.figshare.21876159.v3).

### Statistical analysis

The characteristics of MPO-AAV patients with and without relapse were compared using Fisher’s exact test in two-by-two tables, excluding age, which was compared using the Mann–Whitney U test.

Relapse-free survival curves were generated using the Kaplan-Meier method and were compared among patients who were classified into GPA, MPA, EGPA and unclassifiable (UC) based on the EMA classification, among the patients treated with glucocorticoid (GC) alone, GC plus immunosuppressants, and immunosuppressants alone, and between patients with and without each *HLA class II* allele (*HLA-DRB1, DQA1, DQB1* and *DPB1*), and each amino acid encoded by these *HLA* alleles, using log-rank test. Unadjusted hazard ratios (HRs) and adjusted HRs for EMA classification, treatment (GC alone, GC plus immunosuppressants or immunosuppressants alone), age and sex were calculated using Cox proportional hazard model.

The association of *HLA-DQA1*03:02* and *DRB1*09:01* with susceptibility to MPO-AAV and MPA was tested using logistic regression analysis with an additive model.

Statistical analyses were performed using the R software version 3.5.2. Correction for multiple testing in risk of relapse analysis at each *HLA* locus and amino acid position was performed by controlling the false discovery rate. Q<0.1 was considered significant. There were 17 *DRB1* alleles, 11 *DQA1* alleles, 11*DQB1* alleles, 8 *DPB1* alleles, 79 DR*β*1 amino acids, 47 DQ*α*1 amino acids, 71 DQ*β*1 amino acids, and 26 DP*β*1 amino acids. The log-rank test for the pairwise comparison between the combination of presence/absence of *DQA1*03:02* and DRβ1_13S was corrected for six comparisons, namely, the combination of 2 out of 4 (_4_C_2_=6).

### Ethics

This study was reviewed and approved by the Faculty of Medicine Ethics Committee of University of Tsukuba (Approval ID: 122, 123, 180, 227, 268).

This study was also approved by the Ethics Committees of the following institutes that participated in the collaboration and/or recruitment of subjects: Aichi Medical University, Asahikawa Medical University, Ehime University, Fukuoka University, Hamamatsu University, Hokkaido University, Iwate Prefectural Central Hospital, Juntendo University, Kagawa University. Kanazawa University, Kitano Hospital, Kyorin University, Kyoto University, Kyushu University, Nagasaki University, Nagoya City University, Nagoya University, Nara Medical University, National Defense Medical College, Okayama University, Okayama Saiseikai General Hospital, Saga University, Saitama Medical Center Hospital, Shimane University, The University of Miyazaki, The University of Tokyo, Toho University, Tokyo Medical and Dental University, Tokyo Medical University Hachioji Medical Center, Tokyo Metropolitan Geriatric Hospital, and Institute of Gerontology, and Tokyo Women’s Medical University.

This study was conducted in accordance with the principles of the Declaration of Helsinki and Ethical Guidelines for Human Genome/Gene Analysis Research implemented by the Ministry of Education, Culture, Sports, Science and Technology, the Ministry of Health, Labour and Welfare, and the Ministry of Economy, Trade and Industry, of Japan. Written informed consent was obtained from all the participants.

## Results

### Association of *HLA-DRB1*09:01-DQA1*03:02-DQB1*03:03* haplotype with susceptibility to MPO-AAV and MPA in the Japanese population

We previously reported that *DRB1*09:01* and *DQB1*03:03* are associated with susceptibility to, and *DRB1*13:02* is associated with protection from, MPO-AAV and MPA in a Japanese population (12-15). Recently, *HLA-DQA1*03:02*, which is in strong linkage disequilibrium with *DRB1*09:01* and *DQB1*03:03*, was reported to be associated with MPO-AAV in a Chinese population (16). Because the association of *DQA1*03:02* with MPO-AAV and its relationship with the *DRB1*09:01*-*DQB1*03:03* haplotype have not been previously analyzed in a Japanese population, we addressed this issue.

To genotype *HLA-DQA1*03:02*, we used rs11545686 as a tag single nucleotide variant (tagSNV). The rs11545686 encodes a p.Met18Thr substitution within the signal peptide of the HLA-DQ alpha chain. The rs11545686C allele is carried by *DQA1*03:02, *03:07, *03:13* and *\*03:18* based on the IPD-IMGT/HLA Database (Release 3.47.0, https://www.ebi.ac.uk/ipd/imgt/hla/). Except for *DQA1*03:02*, these alleles are extremely rare in the Japanese population. In fact, rs12722040, which was previously reported to tag *DQA1*03:02* (24), is identical to rs11545686. We confirmed that the *DQA1*03:02* genotyping system based on rs11545686 was concordant with the results of high-resolution allele typing using PCR-SSOP in 199 samples, except for one sample in which the *DQA1* genotype could not be determined using PCR-SSOP.

Using this genotyping system, we examined the association between *DQA1*03:02*, MPO-AAV, and MPA. *DQA1*03:02* was significantly associated with susceptibility to MPO-AAV (P_unconditional_=5.8×10^−7^) and MPA (P_unconditional_=1.1×10^−5^) (Table 2), which confirmed the recent report on a Chinese population (16).

**Table 2.** Association of *DQA1*03:02* with susceptibility to MPO-ANCA positive AAV and MPA

| | n (allele frequency: %) | | $P_{\text{unconditional}}$ / OR (95%CI) | $P_{\text{conditional}}$ / OR (95%CI)* on: | |
| --- | --- | --- | --- | --- | --- |
|  | case | healthy controls |  | *DRB1\*09:01* | *DQA1\*03:02* |
| MPO-AAV | 2n=856 | 2n=1558 |  |  |  |
| *DRB1\*09:01* | 190 (22.2) | 222 (14.2) | $6.9 \times 10^{-7}$ / 1.75 (1.40-2.19) | NA | 0.45 / 1.29 (0.67-2.53) |
| *DQA1\*03:02* | 202 (23.6) | 240 (15.4) | $5.8 \times 10^{-7}$ / 1.74 (1.40-2.16) | 0.33 / 1.37 (0.71-2.61) | NA |
| MPA | 2n=610 | 2n=1558 |  |  |  |
| *DRB1\*09:01* | 137 (22.5) | 222 (14.2) | $4.2 \times 10^{-6}$ / 1.77 (1.39-2.26) | NA | 0.19 / 1.68 (0.79-3.83) |
| *DQA1\*03:02* | 143 (23.4) | 240 (15.4) | $1.1 \times 10^{-5}$ / 1.71 (1.34-2.17) | 0.89 / 1.05 (0.47-2.20) | NA |

OR: odds ratio, CI: confidence interval, MPO-AAV: MPO-ANCA positive ANCA-associated vasculitis, MPA: microscopic polyangiitis, NA: not applicable.
\* $P_{\text{conditional}}$ , OR and 95% CI were conditional on *DRB1\*09:01* or *DQA1\*03:02* in conditional logistic regression analysis.

Strong linkage disequilibrium was observed among *DRB1*09:01, DQA1*03:02* and *DQB1*03:03* (*r*^*2*^: 0.92-0.97). When the associations were conditioned, none remained significant (Table 2). Therefore, we could not determine the single causative allele among *DRB1*09:01, DQA1*03:02* and *DQB1*03:03*.

### Association of *HLA class II* alleles with relapse-free survival in MPO-AAV

Figure 1 shows a flow chart of the MPO-AAV patients enrolled in the relapse-free survival analysis. Among the 264 AAV patients who enrolled in the cohort studies RemIT-JAV (19) and RemIT-JAV-RPGN (20), more than 80% of patients were MPO-ANCA single-positive. As the frequency of relapse was reported to be higher in AAV patients with PR3-ANCA than in those without it (5), this study focused on 199 MPO-ANCA single-positive patients who achieved remission during the observation period. Patient characteristics are shown in Table 3. During the first 3 months after treatment initiation, 84 (42.2%) patients received GC alone, 112 (56.3%) were treated with GC plus immunosuppressants, and three were treated with immunosuppressants alone (1.5%). In most patients, cyclophosphamide was administered as an the immunosuppressant.

**Figure 1.**
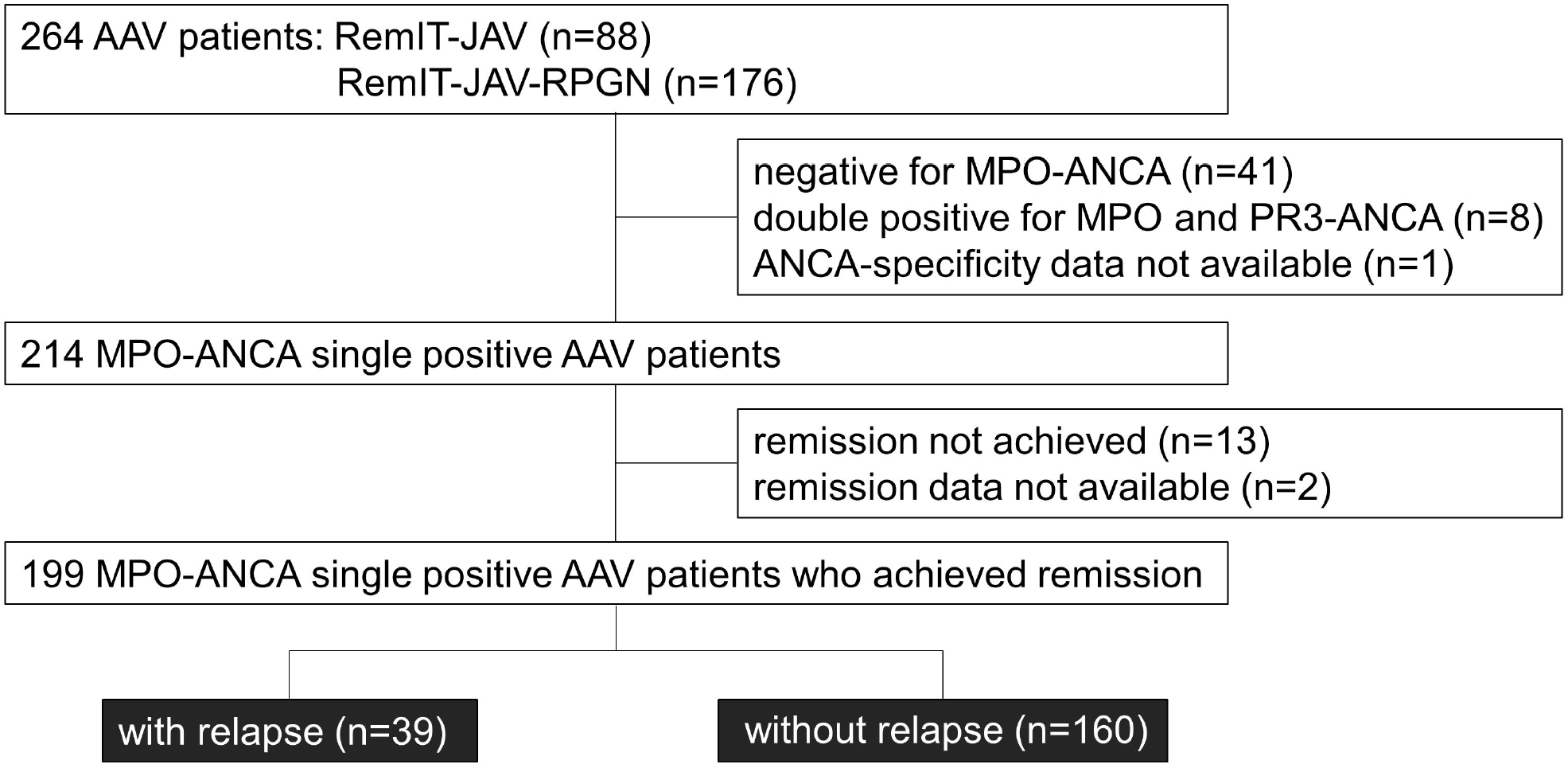
Flow chart of the MPO-AAV patients in the relapse-free survival analysis. Among the patients who entered the cohort studies of remission induction therapy (RemIT-JAV and RemIT-JAV-RPGN), 199 MPO-ANCA positive and PR3-ANCA negative (“MPO-ANCA single-positive”) patients who achieved remission were studied in the relapse-free survival analysis.

**Table 3.**
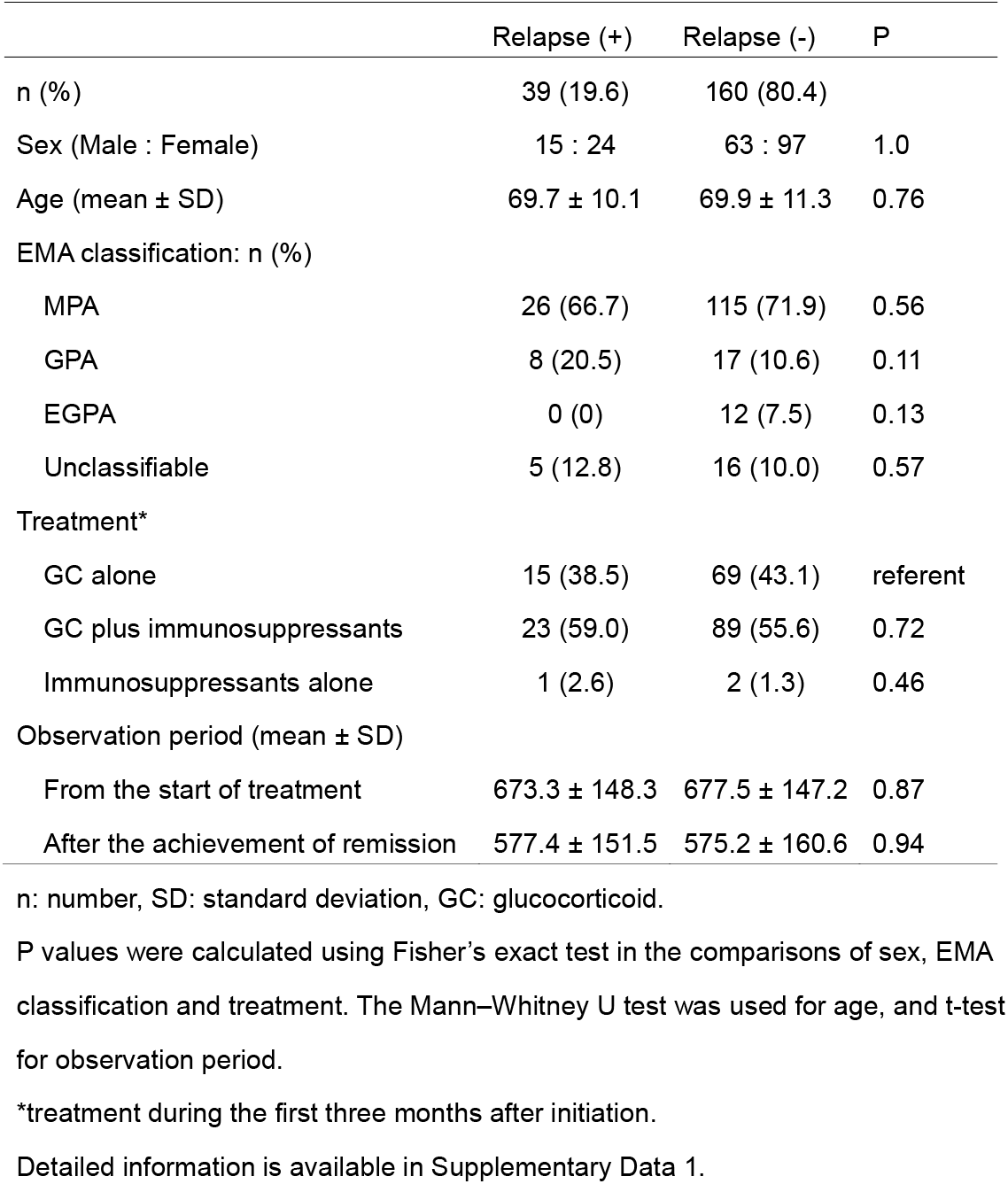
Characteristics of the MPO-ANCA single-positive AAV with and without relapse

The mean observation period from the start of treatment was 676.7 (SD: 147.0) days. Among the 199 MPO-ANCA single positive AAV patients, 39 patients experienced relapse (19.6%). The mean time to relapse after remission was 228.1 (SD: 173.5) days. No significant difference in sex ratio, age, EMA clinical classification, the treatment received, or observation period were observed between patients with and without relapse (Table 3).

Next, we next tested whether the EMA classification and the treatment modalities were associated with the risk of relapse using the Kaplan-Meier method for relapse-free survival. GPA patients have previously been shown to be associated with high occurrence of relapse (5). No significant difference in relapse-free survival was observed among MPO-ANCA single positive AAV patients classified by the EMA algorithm (log-rank test uncorrected P [P_uncorr_]=0.097) (Figure 2A) or treatment modality (log-rank test P_uncorr_=0.78)(Figure 2B).

**Figure 2.**
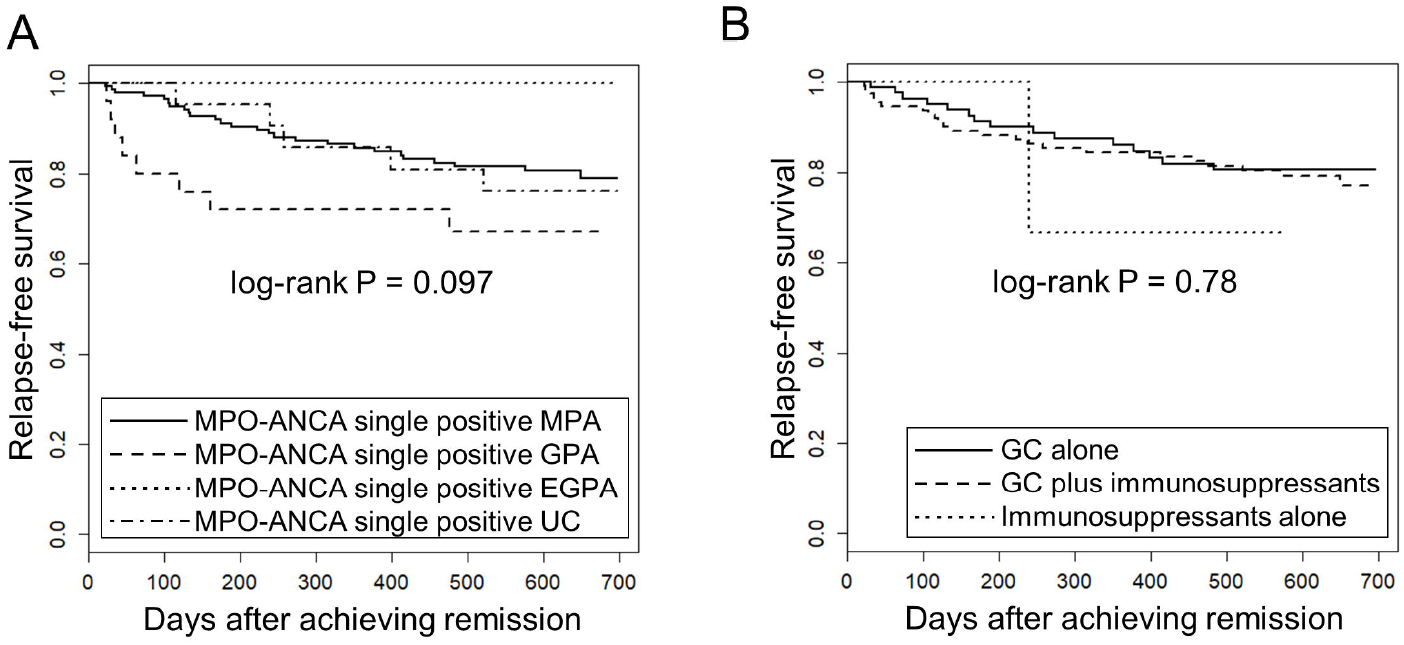
Relapse-free survival in MPO-ANCA single-positive AAV patients according to the EMA classification and treatment modality. The longitudinal and horizontal axes represent the probability of relapse-free survival and days after achievement of remission, respectively. P value was calculated by log-rank test. (A) Relapse-free survival was compared among MPO-ANCA single-positive, MPA (n=141), GPA (n=25), EGPA (n=12) and unclassifiable AAV (UC) (n=21). Note that PR3-ANCA positive patients are not included in any group. (B) Relapse-free survival was compared among MPO-ANCA single-positive AAV patients treated with glucocorticoid (GC) alone (n=84), GC plus immunosuppressants (n=112) and immunosuppressants alone (n=3).

The carrier frequencies of the *HLA-DRB1, DQA1, DQB1* and *DPB1* alleles in MPO-AAV patients with and without relapse are shown in Supplementary Tables S1=S4. Carrier frequencies of *DRB1*09:01* (P_uncorr_=0.049, Q=0.47, odds ratio [OR]: 2.05, 95% confidence interval [CI]: 1.01=4.15), *DQA1*03:02* (P_uncorr_=0.020, Q=0.22, OR: 2.38, 95%CI: 1.16=4.87) and *DQB1*03:03* (P_uncorr_=0.048, Q=0.53, OR: 2.10, 95%CI: 1.03=4.28) were nominally increased in patients with relapse (Table 4). With respect to *DPB1* alleles, no trend toward association was observed among patients with MPO-ANCA single=positive AAV.

**Table 4.**
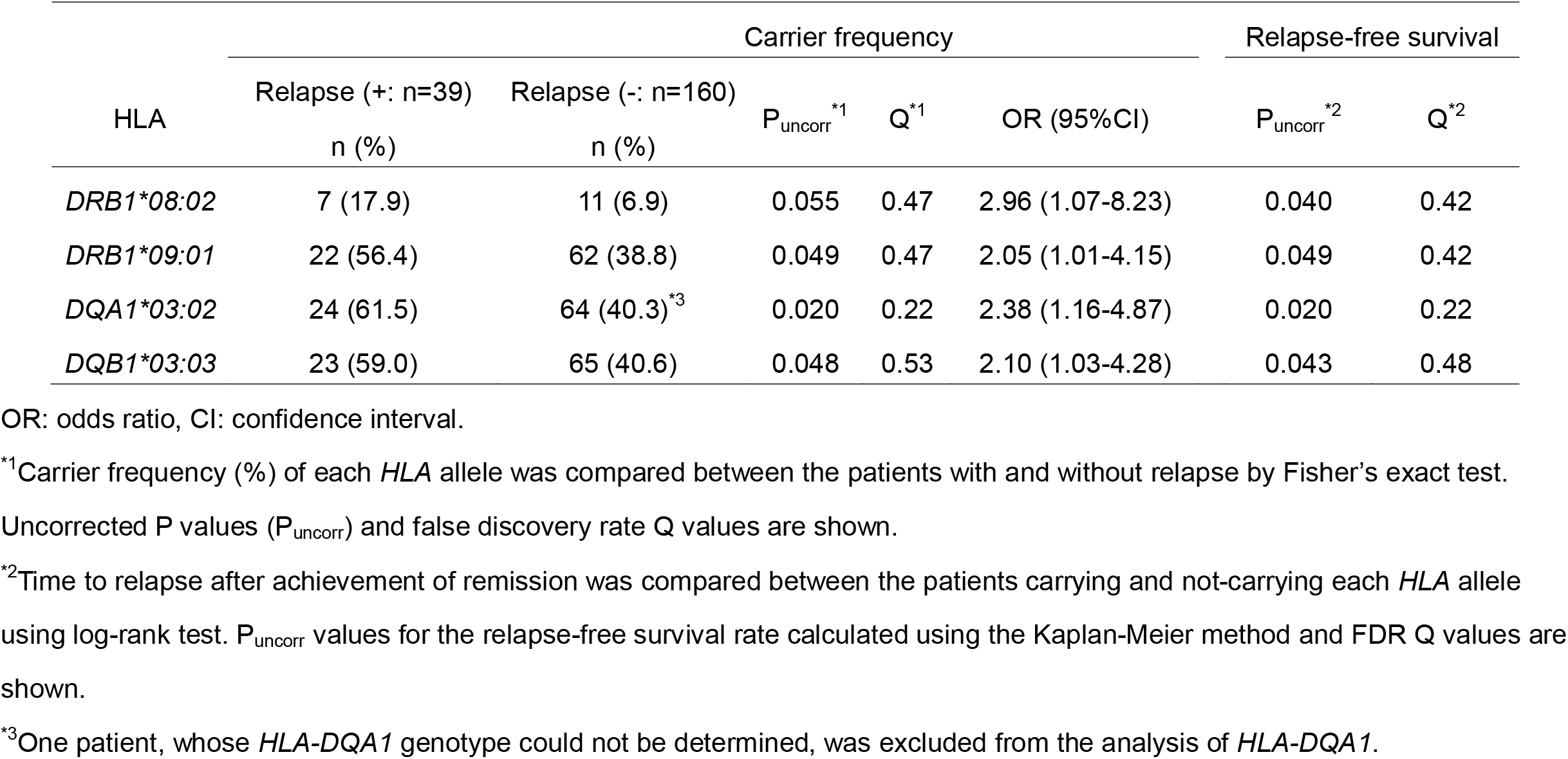
Association of *HLA class II* alleles with occurrence of relapse and risk of relapse

| HLA | Carrier frequency |  |  |  |  | Relapse-free survival |  |
| --- | --- | --- | --- | --- | --- | --- | --- |
|  | Relapse (+: n=39) | Relapse (-: n=160) | P<sub>uncorr</sub><sup>*1</sup> | Q<sup>*1</sup> | OR (95%CI) | P<sub>uncorr</sub><sup>*2</sup> | Q<sup>*2</sup> |
|  | n (%) | n (%) |  |  |  |  |  |
| *DRB1\*08:02* | 7 (17.9) | 11 (6.9) | 0.055 | 0.47 | 2.96 (1.07-8.23) | 0.040 | 0.42 |
| *DRB1\*09:01* | 22 (56.4) | 62 (38.8) | 0.049 | 0.47 | 2.05 (1.01-4.15) | 0.049 | 0.42 |
| *DQA1\*03:02* | 24 (61.5) | 64 (40.3)<sup>*3</sup> | 0.020 | 0.22 | 2.38 (1.16-4.87) | 0.020 | 0.22 |
| *DQB1\*03:03* | 23 (59.0) | 65 (40.6) | 0.048 | 0.53 | 2.10 (1.03-4.28) | 0.043 | 0.48 |

OR: odds ratio, CI: confidence interval.
<sup>\*1</sup>Carrier frequency (%) of each *HLA* allele was compared between the patients with and without relapse by Fisher's exact test.
Uncorrected P values (P<sub>uncorr</sub>) and false discovery rate Q values are shown.
<sup>\*2</sup>Time to relapse after achievement of remission was compared between the patients carrying and not-carrying each *HLA* allele using log-rank test. P<sub>uncorr</sub> values for the relapse-free survival rate calculated using the Kaplan-Meier method and FDR Q values are shown.
<sup>\*3</sup>One patient, whose *HLA-DQA1* genotype could not be determined, was excluded from the analysis of *HLA-DQA1*.

Next, we used the Kaplan-Meier method to compare relapse-free survival between the carriers and non-carriers of each *HLA* allele. As shown in Supplementary Tables S1-S4(https://doi.org/10.6084/m9.figshare.21876159.v3), a nominal association was observed in *DQA1*03:02* (P_uncorr_=0.020, Q=0.22), *DRB1*09:01* (P_uncorr_=0.049, Q=0.42) and *DQB1*03:03* (P_uncorr_=0.043, Q=0.48) which were in linkage disequilibrium with *DQA1*03:02*. In addition, *DRB1*08:02* (P_uncorr_=0.040, Q=0.42) was nominally associated with relapse.

Relapse-free survival curves and rates at the end of the observation period for nominally associated *HLA* alleles are shown in Figure 3A-3D and Table 5. The HRs for relapse were 2.11 (95% CI: 1.11-4.02) in *DQA1*03:02*, 1.87 (0.99-3.52) in *DRB1*09:01*, 1.91 (1.01-3.61) in *DQB1*03:03* and 2.30 (1.01-5.20) in *DRB1*08:02*. No violation of the proportional hazards assumption was observed for these alleles (P>0.05).

**Figure 3.**
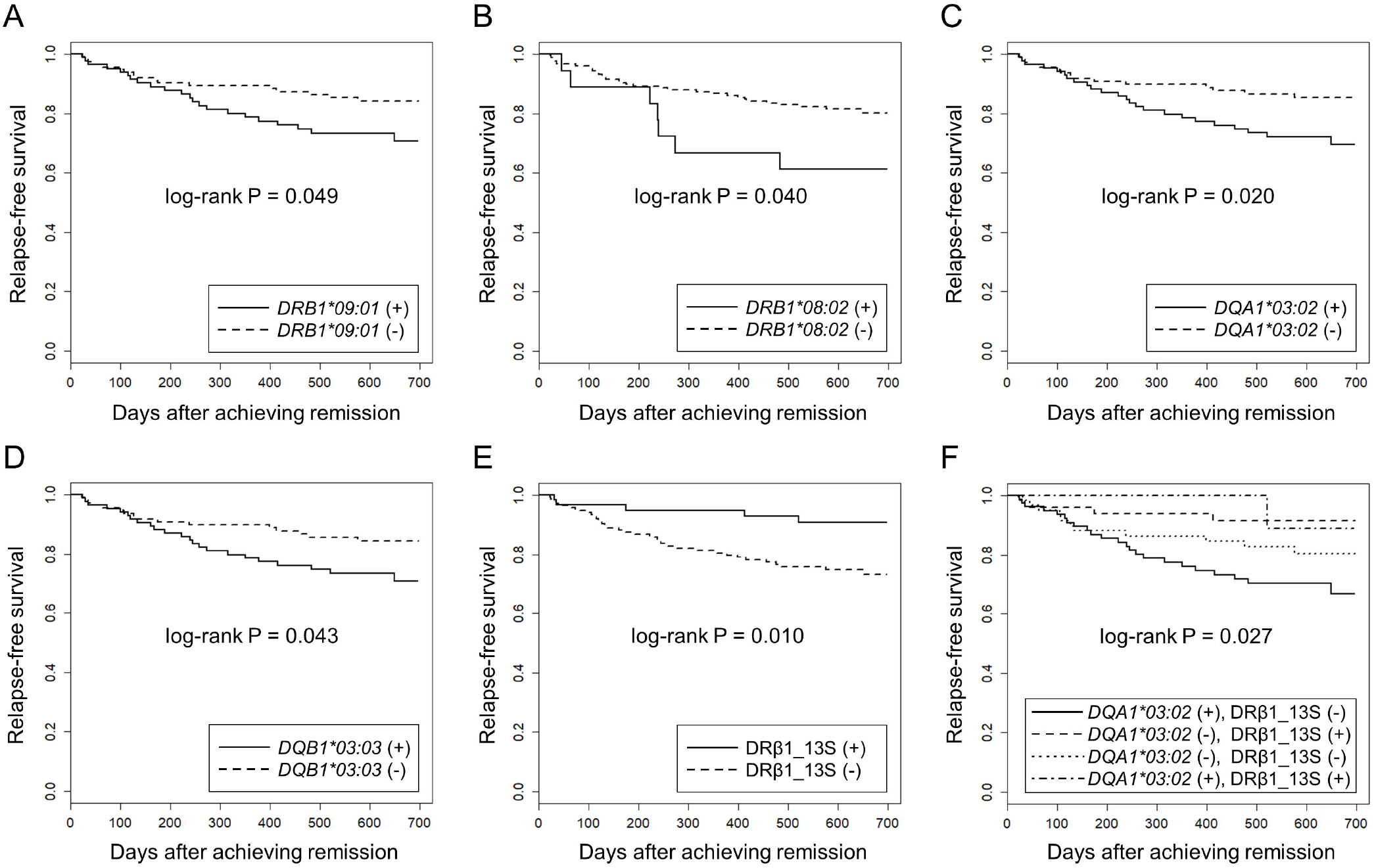
Association of *HLA* alleles and DRβ1 position 13 serine with relapse-free survival in MPO-ANCA single positive AAV patients using Kaplan-Meier method. The longitudinal and horizontal axes represent the probability of relapse-free survival and days after achievement of remission, respectively. Uncorrected P values calculated by log-rank test are shown. Q values corrected for multiple comparisons for each locus are shown in the text, Table 4 and Table 5. Relapse-free survival was compared between MPO-ANCA single-positive AAV patients carrying and not carrying **(A)** *HLA*-*DRB1*09:01*, **(B)** *HLA*-*DRB1*08:02*, **(C)** *HLA-DQA1*03:02*, **(D)** *HLA-DQB1*03:03* and **(E)** HLA-DRβ1 position 13 serine (HLA-DRβ1_13S). **(F)** the patients were divided into four groups according to the carriage of *HLA-DQA1*03:02* and HLA-DRβ1_13S. Relapse-free survival was compared among *DQA1*03:02* (+) and DRβ1_13S (-) (n=79), *DQA1*03:02* (-) and DRβ1_13S (+) (n=49), *DQA1*03:02* (-) and DRβ1_13S (-) (n=61) and *DQA1*03:02* (+) and DRβ1_13S (+) (n=9) groups.

**Table 5.** Association of *HLA class II* alleles and DRβ1 position 13 serine with risk of relapse in multivariate Cox proportional hazards model

|  | relapse-free survival rate (95% CI) |  |  | P<sub>conditional</sub> / HR (95%CI)* on: |  |  |
| --- | --- | --- | --- | --- | --- | --- |
|  | positive patients | negative patients | P<sub>unconditional</sub> / HR (95%CI) | MPA | GPA | Unclassifiable |
| HLA-allele |  |  |  |  |  |  |
| DRB1*08:02 | 61.1 (42.3-88.3) | 80.2 (74.1-86.8) | 0.046 / 2.30 (1.01-5.20) | 0.051 / 2.28 (1.00-5.19) | 0.088 / 2.07 (0.90-4.77) | 0.049 / 2.29 (1.00-5.23) |
| DRB1*09:01 | 70.6 (60.6-82.3) | 84.1 (77.4-91.4) | 0.053 / 1.87 (0.99-3.52) | 0.046 / 1.93 (1.01-3.67) | 0.040 / 1.96 (1.03-3.73) | 0.045 / 1.94 (1.02-3.69) |
| DQA1*03:02 | 69.4 (59.5-81.0) | 85.4 (78.7-92.5) | 0.023 / 2.11 (1.11-4.02) | 0.020 / 2.18 (1.13-4.19) | 0.013 / 2.29 (1.19-4.41) | 0.019 / 2.19 (1.14-4.23) |
| DQB1*03:03 | 70.6 (60.8-82.1) | 84.5 (77.7-91.8) | 0.047 / 1.91 (1.01-3.61) | 0.041 / 1.96 (1.03-3.75) | 0.034 / 2.02 (1.06-3.85) | 0.039 / 1.98 (1.03-3.78) |
| HLA-amino acid |  |  |  |  |  |  |
| DRβ1_13S | 90.9 (83.5-98.9) | 73.0 (65.3-81.5) | 0.015 / 0.31 (0.12-0.80) | 0.014 / 0.31 (0.12-0.79) | 0.018 / 0.32 (0.12-0.82) | 0.014 / 0.30 (0.12-0.78) |

HR: hazard ratio, CI: confidence interval, MPA: microscopic polyangiitis, GPA: granulomatosis with polyangiitis.
\* $P_{\text{conditional}}$ , HR and 95% CI conditional on EMA classification (MPA, GPA or unclassifiable), treatment, age and sex in multivariate Cox proportional hazards model. EGPA was not included in variables of the EMA classification, because none of the patients with EGPA experienced relapse.

When HRs for the associated *HLA* alleles were conditioned on the EMA classification (MPA, GPA or UC), treatment modality during the initial three months (GC alone, immunosuppressants alone and GC plus immunosuppressants), age and sex using a multivariate Cox proportional hazards model, the associations of *DRB1*09:01, DQA1*03:02* and *DQB1*03:03* were not affected, whereas the association of *DRB1*08:02* was attenuated after conditioning (Table 5). In this analysis, EGPA was not included in the variables of the EMA classification, because no patients with EGPA experienced relapse.

Next, we examined whether specific amino acids in the HLA molecules were associated with relapse. The results of the log-rank test for each amino acid are presented in Supplementary Data 2 (https://doi.org/10.6084/m9.figshare.21876159.v3). The strongest association trend was observed in serine residue at position 13 of HLA-DRβ1 (HLA-DRβ1_13S: log-rank P_uncorr_=0.010, Q=0.42). The relapse-free survival period was longer in patients carrying HLA-DRβ1_13S (HR: 0.31, 95% CI: 0.12-0.80) (Figure 3E and Table 5).

When the associations of the *HLA* alleles nominally associated with the risk of relapse and HLA-DRβ1_13S were conditioned on each other, a tendency toward association remained in DRβ1_13S (P_conditional_: 0.057 [conditional on *DQA1*03:02*] and 0.040 [conditional on *DQB1*03:03*]) (Table 6).

**Table 6.** Conditional survival analysis of *HLA* alleles and DRβ1 position 13 serine in multivariate Cox proportional hazards model

|  |  | P<sub>conditional</sub> / HR (95%CI)* on: |  |  |  |
| --- | --- | --- | --- | --- | --- |
|  | P<sub>unconditional</sub> /<br>HR (95%CI) | *DRB1\*09:01* | *DQA1\*03:02* | *DQB1\*03:03* | DRβ1_13S |
| *HLA*-allele |  |  |  |  |  |
| *DRB1\*09:01* | 0.053 /<br>1.87 (0.99-3.52) | NA | 0.63 /<br>0.70 (0.16-2.97) | 0.85 /<br>1.18 (0.21-6.72) | NA |
| *DQA1\*03:02* | 0.023 /<br>2.11 (1.11-4.02) | 0.15 /<br>2.95 (0.67-12.9) | NA | 0.052 /<br>7.45 (0.98-56.7) | 0.15 /<br>1.65 (0.84-3.23) |
| *DQB1\*03:03* | 0.047 /<br>1.91 (1.01-3.61) | 0.58 /<br>1.63 (0.28-9.39) | 0.21 /<br>0.27 (0.04-2.04) | NA | 0.24 /<br>1.49 (0.77-2.89) |
| HLA<br>-amino acid |  |  |  |  |  |
| DRβ1_13S | 0.015 /<br>0.31 (0.12-0.80) | NA | 0.057 /<br>0.39 (0.15-1.03) | 0.040 /<br>0.36 (0.14-0.96) | NA |

HR: hazard ratio, CI: confidence interval, NA: not applicable.
\* $P_{\text{conditional}}$ , HR and 95% CI were conditioned on indicated *HLA* alleles or amino acid in multivariate Cox proportional hazards model.

Finally, the patients were divided into four groups according to the carriage a f *DQA1*03:02* (as a representative of *DRB1*09:01-DQA1*03:02-DQB1*03:03* relapse-risk haplotype) and DRβ1_13S (relapse-protective), and relapse-free survival was compared. As shown in Figure 3F, a significant difference in the time to relapse was detected among the four groups (log-rank P=0.027). In the pairwise comparisons, significant difference was observed between the patients with highest risk combination (*DQA1*03:02* positive and DRβ1_13S negative) and with lowest risk for relapse combination (*DQA1*03:02* negative and DRβ1_13S positive), even after correction for multiple testing (HR: 4.02, 95% CI: 1.39-11.6, P_uncorr_=0.0055, Q=0.033).

## Discussion

In this study, we examined whether *HLA class II* alleles and HLA amino acids were associated with relapse in MPO-AAV, using data from two Japanese nationwide cohort studies on remission induction therapy: RemIT-JAV (19) and RemIT-JAV-RPGN (20). We detected that MPO-ANCA single positive AAV patients carrying *HLA-DRB1*09:01, DQA1*03:02* or *DQB1*03:03* had nominally higher risk of relapse as compared with those without these alleles, while AAV patients carrying serine residue at position 13 of HLA-DRβ1 had a trend toward lower risk of relapse. By combining these two factors, significant differences were observed in relapse-free survival among the patient subgroups. To the best of our knowledge, this is the first study to demonstrate the association between gene and amino acid variations and AAV relapse in an East Asian population.

We previously identified an association between *HLA-DRB1*09:01* and *DQB1*03:03* and susceptibility to MPA/MPO-AAV in a Japanese population (12-15). In the present study, we confirmed the association of *DQA1*03:02* with susceptibility to MPO-AAV in a Japanese population, which was recently reported in a Chinese population (16). Owing to the tight linkage disequilibrium among these three *HLA class II* alleles, we could not determine which of the three alleles played a primary causative role.

Currently, the molecular mechanisms underlying the association between *HLA class II* alleles and MPA/MPO-AAV remain unclear. Among the components of the risk haplotype *DRB1*09:01-DQA1*03:02-DQB1*03:03*, the polymorphic amino acids encoded by *DRB1*09:01* and *DQB1*03:03* directly affect antigenic peptide specificity. Because MPO-ANCA has been strongly implicated in MPO-AAV pathogenesis (25), it is possible that specific antigenic peptides derived from human MPO (26) or microbial peptides mimicking human MPO peptides (27) might be preferentially presented by *HLA-DRB1*09:01* and/or *DQB1*03:03* products, although to our knowledge such peptides have not been identified from the patients for *DRB1*09:01* or *DQB1*03:03* products, Conversely, the specific amino acid encoded by *DQA1*03:02*, Asp160, is located in the α2 domain of HLA-DQ molecule, and may be involved in stabilizing HLA class II homodimers, thereby affecting antigenic peptide specificity (16). Thus, it is possible that multiple molecular mechanisms associated with MHC-peptide complex formation and T cell receptor signaling are involved in the genetic association between the risk haplotype and susceptibility to MPO-AAV.

A previous report showed that *HLA-DPB1*04:01*, the susceptibility allele for GPA, was also associated with an increased risk of AAV relapse in the Dutch and German patients (17). In this study, GPA and PR3-AAV were predominant. In contrast, this association was not observed in a Danish population (28). Although the reason for this discrepancy is unclear, differences in *DPB1*04:01* carrier frequency among patients between these studies (77% (17) and 94% (28), respectively) may play a role. A recent study from the United States reported an association between *HLA-DPB1*04:01* and relapse in PR3-AAV, but not in MPO-AAV (18).

With respect to MPO-AAV, a small-scale study in a European population did not detect a significant association between relapse and HLA-A, -B, or -DR antigens (29). In our study, *HLA-DRB1*09:01*, -*DQA1*03:02* and -*DQB1*03:03*, the susceptibility alleles for MPO-AAV in East Asian populations (12-16) showed a trend toward a higher risk of relapse of MPO-AAV in a Japanese population, possibly suggesting that the susceptibility alleles for AAV may also be associated with relapse in each ethnic group.

Unlike one of European study (17), this study did not detect an association between *DPB1*04:01* and the risk of AAV relapse. This discrepancy likely occurred because this study focused on MPO-ANCA positive AAV, whereas in the previous European study, 51.7% of the subjects were positive for PR3-ANCA (17). In our MPO-AAV data, no significant association was detected between *DPB1*04:01* and relapse (Supplementary Table S4, https://doi.org/10.6084/m9.figshare.21876159.v3), which is consistent with a report from the United States (18). This difference may not be explained merely by the low carrier frequency of *DPB1*04:01* in the Japanese population, because *DPB1*04:01* showed a slight tendency toward decrease in patients with relapse (P=0.70, OR: 0.39). In the Japanese population, the protective allele for MPO-AAV susceptibility, *DRB1*13:02*, is in linkage disequilibrium with *DPB1*04:01* (*r*^*2*^=0.44), and the allele frequency of *DPB1*04:01* was also decreased in MPO-AAV (P=2.1×10^−4^, OR: 0.40) due to linkage disequilibrium with *DRB1*13:02* (14). Thus, to evaluate the effect of *DPB1*04:01* in the Japanese population, PR3-ANCA positive AAV patients with sufficient sample sizes should be investigated in the future.

In contrast to the *HLA-DRB1*09:01-DQA1*03:02-DQB1*03:03* haplotype, alleles carrying HLA-DRβ1_13S were found to show a trend toward protection from relapse. HLA-DRβ1_13S is encoded by DR3, DR11, DR13 and DR14. HLA-DRB1 position 13 amino acid is associated with multiple immune system disorders, and was recently shown to be most strongly associated with individual differences in the T cell receptor complementarity determining region 3 (CDR3) repertoire (30). HRs (95% CI) and log-rank P values for relapse-free survival of alleles encoding HLA-DRβ1_13S with minor allele frequency >0.01 in the Japanese population are shown in Supplementary Table S5 (https://doi.org/10.6084/m9.figshare.21876159.v3). These alleles showed a trend toward protection from relapse, although the differences did not reach statistical significance.

As mentioned above, we previously reported that *DRB1*13:02*, encoding HLA-DRβ1_13S, exhibited a protective effect against the MPO-AAV development (14). The data on susceptibility to MPO-AAV with HLA-DRβ1_13S, as well as with each allele encoding 13S, are also presented in Supplementary Table S5 (https://doi.org/10.6084/m9.figshare.21876159.v3). HLA-DRβ1_13S was protective against the development of MPO-AAV (P_uncorr_=0.0029, OR 0.71, 95%CI 0.56-.89). Among the HLA-DRβ1_13S encoding alleles, the protective effect of *DRB1*13:02* (P_uncorr_=6.9×10^−5^, OR 0.45, 95%CI 0.30-0.66) appeared to be more striking as compared with other HLA-DRβ1_13S encoding alleles. On the other hand, in the relapse analysis, HR was not much different between *DRB1*13:02* and other DRβ1_13S encoding alleles. This could suggest a possibility that the protective effect against relapse may be shared by HLA-DRβ1_13S alleles, while protection from disease occurrence may be mainly ascribed to *DRB1*13:02*. Further studies with larger sample sizes are needed to validate this hypothesis.

When the combined effect of *DQA1*03:02* and DRβ1_13S was examined, time to relapse was significantly shorter in the highest risk combination (*DQA1*03:02* positive and DRβ1_13S negative) when compared with the lowest risk combination (*DQA1*03:02* negative and DRβ1_13S positive), even after correction for multiple testing (Q=0.033). The HR was 4.02, which was higher than that for *HLA-DPB1*04:01* in Dutch and American studies (17,18).

In view of our results, it could be hypothesized that susceptibility alleles other than *HLA* may also be associated with the risk of MPO-AAV relapse. To date, no gene other than *HLA* has been established as a susceptibility gene for MPO-AAV in Asian populations. In a European GWAS, *PTPN22* rs2476601 was reported to be associated with GPA/PR3-AAV and MPA/MPO-AAV (10); however, the risk allele of *PTPN22* is almost absent in Asian populations. Recently, the association of a *BACH2* variant was identified using target sequencing (31). According to the Genome Aggregation Database (gnomAD) (https://gnomad.broadinstitute.org/) (32), the risk allele in the *BACH2* gene is not detected in East Asian populations. In a European population, it was reported that -463 A allele (rs2333227) in the *MPO* gene, which is associated with lower expression of MPO, was also associated with the risk of relapse in patients with MPO-AAV (33), which needs to be replicated in independent studies.

This study had several limitations. AAV is a rare disease, with an annual incidence of 22.6/million people in Japan (2). Because of the limited sample size and lack of independent replication cohort, it should be emphasized that this study is of an exploratory nature. For example, the attenuation of a trend toward relapse in *DRB1*08:02* after conditioning (Table 5) could possibly be caused by insufficient detection power (0.589) due to the lower carrier frequencies of *DRB1*08:02* compared to other alleles (Table 4). In fact, the HR of *DRB1*08:02* remained >2.0 even after conditioning (Table 5). Further studies are required to draw a definitive conclusions.

Additionally, because this study was based on observational cohort studies conducted between 2009 and 2016 (19,20), the potential effects of currently available treatments such as rituximab and avacopan could not be addressed. In this study, relapse was defined as the recurrence or new onset of clinical signs and symptoms attributable to active vasculitis, similar to previous studies on Japanese patients (6,23). Although some previous studies employed strict definition, including a requirement for treatment escalation (17), we did not use this definition, because we thought that whether treatment is reinforced or not partly depends on each physician’s decision, and might possibly cause a bias. Finally, the classification of AAV is based on the EMA algorithm (1), and the compatibility of our findings with the 2022 American College of Rheumatology/European Alliance of Associations for Rheumatology Classification Criteria (34-36) needs to be validated. These limitations should be addressed in future studies.

## Conclusion

In the present study, we demonstrated that *DRB1*09:01, DQA1*03:02* and *DQB1*03:03*, susceptibility alleles for MPO-AAV, are also nominally associated with the risk of relapse in MPO-AAV in the Japanese population. In addition, we found that the carriers of HLA-DRβ1 allele with serine residue at position 13 are nominally associated with lower risk of relapse. By combining these two factors, a statistically significant difference in the time to relapse was observed. These findings may be relevant in detecting individuals at high risk for AAV relapse of after remission, and adjusting treatment.

## Contribution to the Field statement

Anti-neutrophil cytoplasmic antibody (ANCA)-associated vasculitis (AAV), a group of rare systemic vasculitis, is classified as myeloperoxidase (MPO)-ANCA positive (MPO-AAV) and proteinase 3 (PR3)-ANCA positive (PR3-AAV) subsets. PR3-AAV accounts for the majority of AAV in the European populations, while MPO-AAV is dominant in the East Asian populations. Different types of human leukocyte antigens (HLA), which play a substantial role in individual’s differences in immune responses, are associated with risk of PR3-AAV and MPO-AAV development.

Disease relapse is a major problem in the management of AAV, and biomarkers associated with the relapse risk constitute unmet needs. HLA type associated with the risk of developing PR3-AAV has also been shown to be associated the relapse risk in European populations, but no study has been reported in MPO-AAV, which is prevalent in East Asian populations.

Here, we report that the HLA types associated with higher and lower risk for the development of MPO-AAV also showed trend for higher and lower relapse risk, respectively. When the risk and protective HLA types are combined, the difference reached statistical significance.

These findings may provide a clue for biomarkers to predict the relapse risk of MPO-AAV, and eventually contribute to establishment of precision medicine. (196 words)

## Data Availability

The datasets supporting the conclusions of this article are included within the article, and Supplementary Table S1-S5, Supplementary Figure S1, Supplementary Data 1 and 2 files available at the URL https://doi.org/10.6084/m9.figshare.21876159.v3. Further inquiries can be directed to the corresponding authors.

## Author Contributions

Dr. Kawasaki and Dr. Tsuchiya designed the study, interpreted the data and wrote the manuscript. Dr. Sada, Dr. Yamagata, Dr. Hashimoto, Dr. Makino, Dr. Arimura and Dr. Harigai coordinated the cohorts. Dr. Kawasaki and Ms. Premita Ari performed genotyping and statistical analyses. Dr. Sada, Dr. Hirano, Dr. Kobayashi, Dr. Nagasaka, Dr. Sugihara, Dr. Ono, Dr. Fujimoto, Dr. Kusaoi, Dr. Tamura, Dr. Kusanagi, Dr. Itoh and Dr. Sumida recruited the participants and collected clinical data. All authors read and approved the final version of the manuscript.

## Acknowledgements

The authors are grateful to all the patients for participating in this study, and to the clinical staff associated with Japan Research Committee of the Ministry of Health, Labour, and Welfare for Intractable Vasculitis (JPVAS) and Research Committee of Intractable Renal Disease of the Ministry of Health, Labour, and Welfare of Japan for recruiting the patients and collecting clinical information. We would like to thank Editage (www.editage.com) for English language editing.

## Figure Legends

**Supplementary Figure S1. Sanger sequencing chromatograms around the *HLA-DQA1*03:02* tagSNV rs11545686**

Representative Sanger sequencing chromatograms corresponding to each of the three genotypes are shown. The rs11545686C allele tags *HLA-DQA1*03:02. HLA-DQA1* genotype determined by PCR-SSOP was *DQA1*03:02/03*02* (relapse ID 36), *DQA1*03:02/05:07* (relapse ID 156) and *DQA1* 01:01/01:02* (relapse ID 164), as shown in Supplementary Data 1.

(available at https://doi.org/10.6084/m9.figshare.21876159.v3)

## References

1. Watts R, Lane S, Hanslik T, Hauser T, Hellmich B, Koldingsnes W, et al. Development and validation of a consensus methodology for the classification of the ANCA-associated vasculitides and polyarteritis nodosa for epidemiological studies. Ann Rheum Dis (2007) 66:222–7. doi:10.1136/ard.2006.054593

2. Fujimoto S, Watts RA, Kobayashi S, Suzuki K, Jayne DR, Scott DG, et al. Comparison of the epidemiology of anti-neutrophil cytoplasmic antibody-associated vasculitis between Japan and the U.K. Rheumatology (2011) 50:1916–20. doi:10.1093/rheumatology/ker205

3. Novikov PI, Smitienko I, Moiseev SV. Duration of maintenance therapy for ANCA-associated vasculitis: more questions than answers. Ann Rheum Dis (2018) 77:e29.doi:10.1136/annrheumdis-2017-211972

4. Pierrot-Deseilligny Despujol C, Pouchot J, Pagnoux C, Pagnoux C, Coste J, Guillevin L, et al. Predictors at diagnosis of a first Wegener’s granulomatosis relapse after obtaining complete remission. Rheumatology (2010) 49:2181–90.doi:10.1093/rheumatology/keq244

5. Terrier B, Pagnoux C, Perrodeau É, Karras A, Khouatra C, Aumaître O, et al. Long-term efficacy of remission-maintenance regimens for ANCA-associated vasculitides. Ann Rheum Dis (2018) 77:1151–7. doi:10.1136/annrheumdis-2017-212768

6. Wada T, Hara A, Arimura Y, Sada KE, Makino H. Risk factors associated with relapse in Japanese patients with microscopic polyangiitis. J Rheumatol (2012) 39:545–51. doi:10.3899/jrheum.110705

7. Hara A, Wada T, Sada KE, Amano K, Dobashi H, Harigai M, et al. Risk factors for relapse of antineutrophil cytoplasmic antibody-associated vasculitis in Japan: A nationwide, prospective cohort study. J Rheumatol (2018) 45:521–8. doi:10.3899/jrheum.170508

8. Lyons PA, Rayner TF, Trivedi S, Holle JU, Watts RA, Jayne DR, et al. Genetically distinct subsets within ANCA-associated vasculitis. N Engl J Med (2012) 367:214–23. doi:10.1056/NEJMoa1108735

9. Xie G, Roshandel D, Sherva R, Monach PA, Lu EY, Kung T, et al. Association of granulomatosis with polyangiitis (Wegener’s) with HLA-DPB1*04 and SEMA6A gene variants: evidence from genome-wide analysis. Arthritis Rheum (2013) 65:2457–68. doi:10.1002/art.38036

10. Merkel PA, Xie G, Monach PA, Ji X, Ciavatta DJ, Byun J, et al. Identification of functional and expression polymorphisms associated with risk for antineutrophil cytoplasmic autoantibody-associated vasculitis. Arthritis Rheumatol (2017) 69:1054–66. doi:10.1002/art.40034

11. Heckmann M, Holle JU, Arning L, Knaup S, Hellmich B, Nothnagel M, et al. The Wegener’s granulomatosis quantitative trait locus on chromosome 6p21.3 as characterised by tagSNP genotyping. Ann Rheum Dis (2008) 67:972–9. doi:10.1136/ard.2007.077693

12. Tsuchiya N, Kobayashi S, Kawasaki A, Kyogoku C, Arimura Y, Yoshida M, et al. Genetic background of Japanese patients with antineutrophil cytoplasmic antibody-associated vasculitis: association of HLA-DRB1*0901 with microscopic polyangiitis. J Rheumatol (2003) 30:1534–40.

13. Tsuchiya N, Kobayashi S, Hashimoto H, Ozaki S, Tokunaga K. Association of HLA-DRB1*0901-DQB1*0303 haplotype with microscopic polyangiitis in Japanese. Genes Immun (2006) 7:81–4. doi: 10.1038/sj.gene.6364262.

14. Kawasaki A, Hasebe N, Hidaka M, Hirano F, Sada KE, Kobayashi S, et al. Protective role of HLA-DRB1*13:02 against microscopic polyangiitis and MPO-ANCA-positive vasculitides in a Japanese population: a case-control study. PLoS One (2016) 11:e0154393. doi:10.1371/journal.pone.0154393

15. Kawasaki A, Tsuchiya N. Advances in the genomics of ANCA-associated vasculitis-a view from East Asia. Genes Immun (2021) 22:1–11. doi: 10.1038/s41435-021-00123-x

16. Wang HY, Cui Z, Pei ZY, Fang SB, Chen SF, Zhu L, et al. Risk HLA class II alleles and amino acid residues in myeloperoxidase-ANCA-associated vasculitis. Kidney Int (2019) 96:1010–9. doi:10.1016/j.kint.2019.06.015

17. Hilhorst M, Arndt F, Kemna MJ, Wieczorek S, Donner Y, Wilde B, et al. HLA-DPB1 as a risk factor for relapse in antineutrophil cytoplasmic antibody-associated vasculitis: a cohort study. Arthritis Rheumatol (2016) 68:1721–30. doi:10.1002/art.39620

18. Chen DP, McInnis EA, Wu EY, Stember KG, Hogan SL, Hu Y, et al. Immunological interaction of HLA-DPB1 and proteinase 3 in ANCA vasculitis is associated with clinical disease activity. J Am Soc Nephrol (2022) 33:1517–27. doi: 10.1681/ASN.2021081142

19. Sada KE, Yamamura M, Harigai M, Fujii T, Dobashi H, Takasaki Y, et al. Classification and characteristics of Japanese patients with antineutrophil cytoplasmic antibody-associated vasculitis in a nationwide, prospective, inception cohort study. Arthritis Res Ther (2014) 16:R101. doi:10.1186/ar4550

20. Sada KE, Harigai M, Amano K, Atsumi T, Fujimoto S, Yuzawa Y, et al. Comparison of severity classification in Japanese patients with antineutrophil cytoplasmic antibody-associated vasculitis in a nationwide, prospective, inception cohort study. Mod Rheumatol (2016) 26:730–7. doi:10.3109/14397595.2016.1140274

21. Flossmann O, Bacon P, de Groot K, Jayne D, Rasmussen N, Seo P, et al. Development of comprehensive disease assessment in systemic vasculitis. Ann Rheum Dis (2007) 66:283–92. doi:10.1136/ard.2005.051078

22. Hellmich B, Flossmann O, Gross WL, Bacon P, Cohen-Tervaert JW, Guillevin L, et al. EULAR recommendations for conducting clinical studies and/or clinical trials in systemic vasculitis: focus on anti-neutrophil cytoplasm antibody-associated vasculitis. Ann Rheum Dis (2007) 66:605–17. doi:10.1136/ard.2006.062711

23. Watanabe H, Sada KE, Matsumoto Y, Harigai M, Amano K, Dobashi H, et al. Association between reappearance of myeloperoxidase-antineutrophil cytoplasmic antibody and relapse in antineutrophil cytoplasmic antibody-associated vasculitis: subgroup analysis of nationwide prospective cohort studies. Arthritis Rheumatol (2018) 70:1626–33. doi:10.1002/art.40538

24. Zhou F, Cao H, Zuo X, Zhang T, Zhang X, Liu X, et al. Deep sequencing of the MHC region in the Chinese population contributes to studies of complex disease. Nat Genet (2016) 48:740–6. doi:10.1038/ng.3576

25. Kitching AR, Anders HJ, Basu N, Brouwer E, Gordon J, Jayne DR, et al. ANCA-associated vasculitis. Nat Rev Dis Primers (2020) 27;6:71. doi: 10.1038/s41572-020-0204-y.

26. Free ME, Stember KG, Hess JJ, McInnis EA, Lardinois O, Hogan SL, et al. Restricted myeloperoxidase epitopes drive the adaptive immune response in MPO-ANCA vasculitis. J Autoimmun (2020) 106:102306. doi: 10.1016/j.jaut.2019.102306.

27. Ooi JD, Jiang JH, Eggenhuizen PJ, Chua LL, van Timmeren M, Loh KL, et al. A plasmid-encoded peptide from Staphylococcus aureus induces anti-myeloperoxidase nephritogenic autoimmunity. Nat Commun (2019) 29;10:3392. doi: 10.1038/s41467-019-11255-0.

28. Gregersen JW, Erikstrup C, Ivarsen P, Glerup R, Krarup E, Keller KK, et al. PR3-ANCA-associated vasculitis is associated with a specific motif in the peptide-binding cleft of HLA-DP molecules. Rheumatology (2019) 58:1942–9. doi:10.1093/rheumatology/kez111

29. Stassen PM, Cohen-Tervaert JW, Lems SP, Hepkema BG, Kallenberg CGM, Stegeman CA, et al. HLA-DR4, DR13(6) and the ancestral haplotype A1B8DR3 are associated with ANCA-associated vasculitis and Wegener’s granulomatosis. Rheumatology (2009) 48:622–5. doi:10.1093/rheumatology/kep057

30. Ishigaki K, Lagattuta KA, Luo Y, James EA, Buckner JH, Raychaudhuri S. HLA autoimmune risk alleles restrict the hypervariable region of T cell receptors. Nat Genet (2022) 54:393–402. doi: 10.1038/s41588-022-01032-z

31. Dahlqvist J, Ekman D, Sennblad B, Kozyrev SV, Nordin J, Karlsson A, et al. Identification and functional characterization of a novel susceptibility locus for small vessel vasculitis with MPO-ANCA. Rheumatology (2022) 61:3461–70. doi: 10.1093/rheumatology/keab912

32. Karczewski KJ, Francioli LC, Tiao G, Cummings BB, Alföldi J, Wang Q, et al. The mutational constraint spectrum quantified from variation in 141,456 humans. Nature (2020) 581:434–43. doi:10.1038/s41586-020-2308-7

33. Reynolds WF, Stegeman CA, Tervaert JW. -463 G/A myeloperoxidase promoter polymorphism is associated with clinical manifestations and the course of disease in MPO-ANCA-associated vasculitis. Clin Immunol (2002) 103:154–60. doi:10.1006/clim.2002.5206

34. Grayson PC, Ponte C, Suppiah R, Robson JC, Craven A, Judge A, et al. 2022 American College of Rheumatology/European Alliance of Associations for Rheumatology classification criteria for eosinophilic granulomatosis with polyangiitis. Ann Rheum Dis (2022) 81:309–14. doi: 10.1136/annrheumdis-2021-221794.

35. Robson JC, Grayson PC, Ponte C, Suppiah R, Craven A, Judge A, et al. 2022 American College of Rheumatology/European Alliance of Associations for Rheumatology classification criteria for granulomatosis with polyangiitis. Ann Rheum Dis (2022) 81:315–20. doi: 10.1136/annrheumdis-2021-221795.

36. Suppiah R, Robson JC, Grayson PC, Ponte C, Craven A, Khalid S, et al. 2022 American College of Rheumatology/European Alliance of Associations for Rheumatology classification criteria for microscopic polyangiitis. Ann Rheum Dis (2022) 81:321–6. doi: 10.1136/annrheumdis-2021-221796

