## Supplementary Tables S1-S5 for "Association of *HLA-class II* alleles with risk of relapse in myeloperoxidase-antineutrophil cytoplasmic antibody positive vasculitis in the Japanese population"

Supplementary Table S1. Association analysis of each *HLA-DRB1* allele with occurrence of relapse and time to relapse

| | Carrier frequency | | | | Relapse-free survival |
|---|---|---|---|---|---|
| | Relapse (+: n=39) n (%) | Relapse (-: n=160) n (%) | $P_{uncorr}$*1 | OR (95%CI) | log-rank $P_{uncorr}$*2 |
| *DRB1*01:01* | 5 (12.8) | 28 (17.5) | 0.63 | 0.69 (0.25-1.93) | 0.53 |
| *DRB1*04:01* | 1 (2.6) | 1 (0.6) | 0.35 | 4.18 (0.26-68.4) | 0.37 |
| *DRB1*04:05* | 7 (17.9) | 35 (21.9) | 0.67 | 0.78 (0.32-1.92) | 0.58 |
| *DRB1*04:06* | 4 (10.3) | 8 (5.0) | 0.26 | 2.17 (0.62-7.62) | 0.25 |
| *DRB1*04:10* | 1 (2.6) | 3 (1.9) | 0.59 | 1.38 (0.14-13.6) | 0.65 |
| *DRB1*08:02**3 | 7 (17.9) | 11 (6.9) | 0.055 | 2.96 (1.07-8.23) | 0.040 |
| *DRB1*08:03* | 4 (10.3) | 15 (9.4) | 0.77 | 1.10 (0.35-3.53) | 0.83 |
| *DRB1*09:01**3 | 22 (56.4) | 62 (38.8) | 0.049 | 2.05 (1.01-4.15) | 0.049 |
| *DRB1*11:01* | 1 (2.6) | 8 (5.0) | 1.00 | 0.50 (0.06-4.12) | 0.47 |
| *DRB1*12:01* | 3 (7.7) | 11 (6.9) | 0.74 | 1.13 (0.30-4.26) | 0.87 |
| *DRB1*12:02* | 1 (2.6) | 3 (1.9) | 0.59 | 1.38 (0.14-13.6) | 0.58 |
| *DRB1*13:02* | 1 (2.6) | 14 (8.8) | 0.31 | 0.27 (0.03-2.15) | 0.22 |
| *DRB1*14:05* | 1 (2.6) | 5 (3.1) | 1.00 | 0.82 (0.09-7.19) | 0.79 |
| *DRB1*14*54* | 3 (7.7) | 15 (9.4) | 1.00 | 0.81 (0.22-2.93) | 0.79 |
| *DRB1*15:01* | 5 (12.8) | 16 (10.0) | 0.57 | 1.32 (0.45-3.86) | 0.60 |
| *DRB1*15:02* | 9 (23.1) | 34 (21.3) | 0.83 | 1.11 (0.48-2.56) | 0.80 |
| *DRB1*16:02* | 1 (2.6) | 3 (1.9) | 0.59 | 1.38 (0.14-13.6) | 0.74 |

OR: odds ratio, CI: confidence interval.

*1Carrier frequency (%) of each *HLA-DRB1* allele was compared between the patients with and without relapse by Fisher's exact test. Uncorrected P values ($P_{uncorr}$) are shown.

*2Time to relapse after achievement of remission was compared between the patients carrying and not-carrying each *HLA-DRB1* allele. Log-rank $P_{uncorr}$ values for the relapse-free survival rate calculated by Kaplan-Meier method are shown.

*3For *DRB1*08:02* and *DRB1*09:01*, FDR Q values are shown in the main document (Table 4).

Supplementary Table S2. Association analysis of each *HLA-DQA1* allele with occurrence of relapse and time to relapse

| | Carrier frequency | | | | Relapse-free survival |
|---|---|---|---|---|---|
| | Relapse (+: n=39) n (%) | Relapse (-: n=159) n (%) | $P_{uncorr}$ *1 | OR (95%CI) | log-rank $P_{uncorr}$ *2 |
| *DQA1*01:01* | 5 (12.8) | 28 (17.6) | 0.63 | 0.69 (0.25-1.91) | 0.52 |
| *DQA1*01:02* | 7 (17.9) | 30 (18.9) | 1.0 | 0.94 (0.38-2.33) | 0.96 |
| *DQA1*01:03* | 13 (33.3) | 47 (29.6) | 0.70 | 1.19 (0.56-2.52) | 0.65 |
| *DQA1*01:04* | 3 (7.7) | 21 (13.2) | 0.42 | 0.55 (0.15-1.94) | 0.38 |
| *DQA1*03:01* | 7 (17.9) | 27 (17) | 1.0 | 1.07 (0.43-2.67) | 0.94 |
| *DQA1*03:02* *3 | 24 (61.5) | 64 (40.3) | 0.020 | 2.38 (1.16-4.87) | 0.020 |
| *DQA1*03:03* | 9 (23.1) | 37 (23.3) | 1.0 | 0.99 (0.43-2.27) | 0.97 |
| *DQA1*04:01* | 4 (10.3) | 5 (3.1) | 0.077 | 3.52 (0.90-13.8) | 0.075 |
| *DQA1*05:05* | 1 (2.6) | 14 (8.8) | 0.31 | 0.27 (0.03-2.14) | 0.18 |
| *DQA1*05:08* | 2 (5.1) | 3 (1.9) | 0.26 | 2.81 (0.45-17.4) | 0.18 |
| *DQA1*06:01* | 1 (2.6) | 4 (2.5) | 1.0 | 1.02 (0.11-9.39) | 0.70 |

OR: odds ratio, CI: confidence interval.

*1 Carrier frequency (%) of each *HLA-DQA1* allele was compared between the patients with and without relapse by Fisher's exact test. Uncorrected P values ($P_{uncorr}$) are shown.

*2 Time to relapse after achievement of remission was compared between the patients carrying and not-carrying each *HLA-DQA1* allele. Log-rank $P_{uncorr}$ values for the relapse-free survival rate calculated by Kaplan-Meier method are shown.

[*3] For *DQA1*03:02*, FDR Q value is shown in the main document (Table 4).

Supplementary Table S3. Association analysis of each *HLA-DQB1* allele with occurrence of relapse and time to relapse

| | Carrier frequency | | | | Relapse-free survival |
|---|---|---|---|---|---|
| | Relapse (+: n=39) n (%) | Relapse (-: n=160) n (%) | $P_{uncorr}$ [*1] | OR (95%CI) | log-rank $P_{uncorr}$ [*2] |
| *DQB1*03:01* | 5 (12.8) | 36 (22.5) | 0.27 | 0.51 (0.18-1.39) | 0.21 |
| *DQB1*03:02* | 7 (17.9) | 27 (16.9) | 0.82 | 1.08 (0.43-2.69) | 0.93 |
| *DQB1*03:03*[*3] | 23 (59.0) | 65 (40.6) | 0.048 | 2.10 (1.03-4.28) | 0.043 |
| *DQB1*04:01* | 7 (17.9) | 34 (21.3) | 0.83 | 0.81 (0.33-2.00) | 0.63 |
| *DQB1*04:02* | 5 (12.8) | 10 (6.3) | 0.18 | 2.21 (0.71-6.87) | 0.18 |
| *DQB1*05:01* | 5 (12.8) | 28 (17.5) | 0.63 | 0.69 (0.25-1.93) | 0.53 |
| *DQB1*05:02* | 2 (5.1) | 12 (7.5) | 1.0 | 0.67 (0.14-3.11) | 0.65 |
| *DQB1*05:03* | 2 (5.1) | 12 (7.5) | 1.0 | 0.67 (0.14-3.11) | 0.63 |
| *DQB1*06:01* | 13 (33.3) | 47 (29.4) | 0.70 | 1.20 (0.57-2.54) | 0.63 |
| *DQB1*06:02* | 5 (12.8) | 15 (9.4) | 0.55 | 1.42 (0.48-4.18) | 0.50 |
| *DQB1*06:04* | 1 (2.6) | 13 (8.1) | 0.31 | 0.30 (0.04-2.35) | 0.26 |

OR: odds ratio, CI: confidence interval.

[*1]Carrier frequency (%) of each *HLA-DQB1* allele was compared between the patients with and without relapse by Fisher's exact test. Uncorrected P values ($P_{uncorr}$) are shown.

[*2]Time to relapse after achievement of remission was compared between the patients carrying and not-carrying each *HLA-DQB1* allele. Log-rank $P_{uncorr}$ values for the relapse-free survival rate calculated by Kaplan-Meier method are shown.

[*3]For *DQB1*03:03*, FDR Q value is shown in the main document (Table 4).

Supplementary Table S4. Association analysis of each *HLA-DPB1* allele with occurrence of relapse and time to relapse

| | Carrier frequency | | | | Relapse-free survival |
|---|---|---|---|---|---|
| | Relapse (+: n=39) n (%) | Relapse (-: n=160) n (%) | $P_{uncorr}$ *1 | OR (95%CI) | log-rank $P_{uncorr}$ *2 |
| *DPB1*02:01* | 22 (56.4) | 72 (45.0) | 0.22 | 1.58 (0.78-3.20) | 0.27 |
| *DPB1*02:02* | 2 (5.1) | 13 (8.1) | 0.74 | 0.61 (0.13-2.83) | 0.58 |
| *DPB1*03:01* | 5 (12.8) | 14 (8.8) | 0.54 | 1.53 (0.52-4.55) | 0.38 |
| *DPB1*04:01* | 1 (2.6) | 10 (6.3) | 0.70 | 0.39 (0.05-3.18) | 0.41 |
| *DPB1*04:02* | 8 (20.5) | 42 (26.3) | 0.54 | 0.73 (0.31-1.70) | 0.54 |
| *DPB1*05:01* | 22 (56.4) | 87(54.4) | 0.86 | 1.09 (0.54-2.20) | 0.81 |
| *DPB1*09:01* | 8 (20.5) | 31 (19.4) | 0.83 | 1.07 (0.45-2.56) | 0.89 |
| *DPB1*13:01* | 2 (5.1) | 4 (2.5) | 0.33 | 2.11 (0.37-11.9) | 0.45 |

OR: odds ratio, CI: confidence interval.

*1Carrier frequency (%) of each *HLA-DPB1* allele was compared between the patients with and without relapse by Fisher's exact test. Uncorrected P values ($P_{uncorr}$) are shown.

*2Time to relapse after achievement of remission was compared between the patients carrying and not-carrying each *HLA-DPB1* allele. Log-rank $P_{uncorr}$ values for the relapse-free survival rate calculated by Kaplan-Meier method are shown.

Supplementary Table S5 Association analyses of HLA-DRβ1_13S encoding alleles with susceptibility and relapse-free survival.

| | susceptibility | | | | relapse-free survival | |
|---|---|---|---|---|---|---|
| | MPO-AAV (2n=856) | healthy controls (2n=1558) | $P_{uncorr}$ | OR / 95% CI | log-rank $P_{uncorr}$ | HR / 95%CI |
| HLA amino acid | | | | | | |
| DRβ1_13S | 130 (15.2) | 312 (20.0) | 0.0029 | 0.71 / 0.56-0.89 | 0.015 | 0.31 / 0.12-0.80 |
| *HLA*-allele encoding DRβ1_13S | | | | | | |
| *DRB1*11:01* | 24 (2.8) | 30 (1.9) | 0.16 | 1.48 / 0.85-2.57 | 0.47 | 0.49 / 0.07-3.55 |
| *DRB1*13:02* | 32 (3.7) | 126 (8.1) | $6.9x10^{-5}$ | 0.45 / 0.30-0.66 | 0.22 | 0.31 / 0.04-2.24 |
| *DRB1*14:03* | 22 (2.6) | 32 (2.1) | 0.41 | 1.26 / 0.72-2.19 | - | - |
| *DRB1*14:05* | 15 (1.8) | 48 (3.1) | 0.054 | 0.56 / 0.30-0.98 | 0.79 | 0.77 / 0.11-5.59 |
| *DRB1*14:06* | 5 (0.6) | 16 (1.0) | 0.27 | 0.56 / 0.18-1.45 | - | - |
| *DRB1*14:54* | 31 (3.6) | 50 (3.2) | 0.59 | 1.13 / 0.71-1.78 | 0.79 | 0.85 / 0.26-2.76 |

Associations with susceptibility and relapse-free survival of HLA-DRβ1_13S encoding alleles in the subjects of this study. Allele frequencies are shown in parentheses. Association with susceptibility was examined using logistic regression analysis under the additive model. Only the alleles with minor allele frequency >0.01 are listed.

The data on relapse-free survival are derived from Table 5 (main text) and Supplementary Table S1. Carriers of *DRB1*14:03* and *DRB1*14:06* were not present in the subjects with relapse.

OR: odds ratio, CI: confidence interval, HR: hazard ratio.
