## Supplementary figures and images for "Association of *HLA-class II* alleles with risk of relapse in myeloperoxidase-antineutrophil cytoplasmic antibody positive vasculitis in the Japanese population"

### Supplementary Figure S1

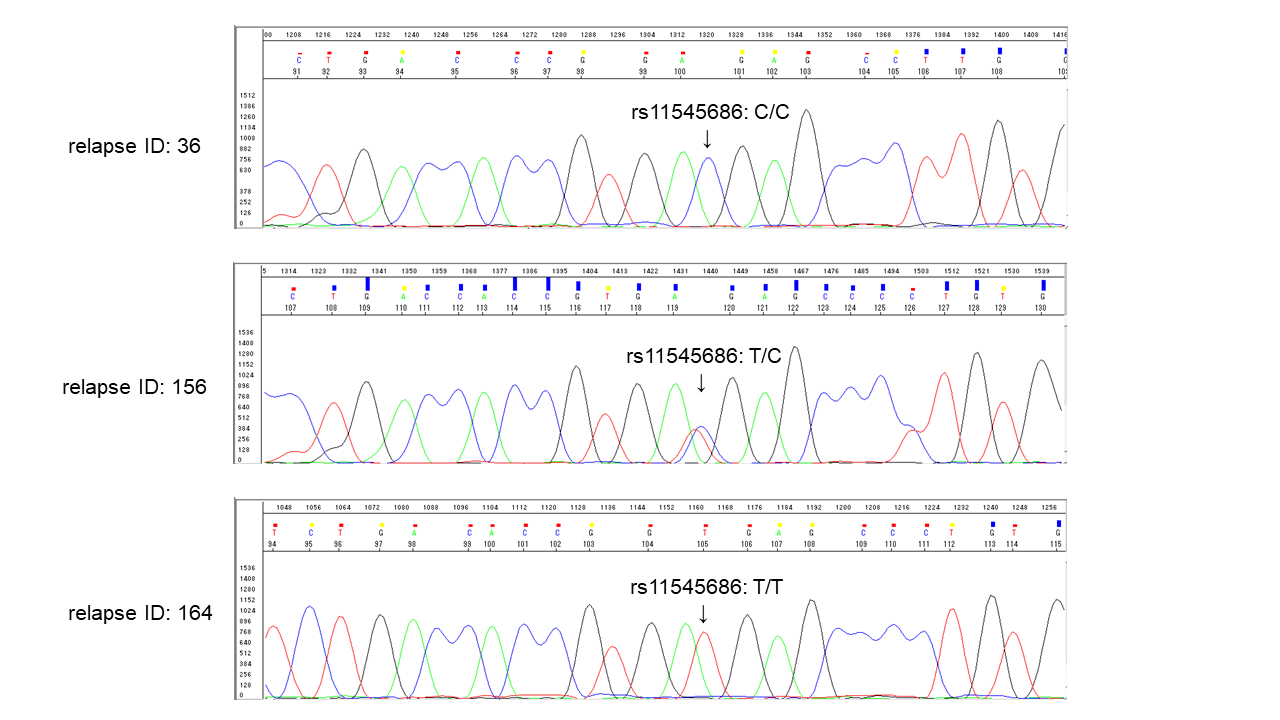
